# Prospective screening of liver fibrosis in a primary care cohort using systematic calculation of fib-4 in routine results

**DOI:** 10.1101/2021.01.13.21249262

**Authors:** Philippe Halfon, Christelle Ansaldi, Guillaume Penaranda, Laurent Chiche, Patrick Dukan, Chloé Stavris, Anne Plauzolles, Frédérique Retornaz, Marc Bourliere

## Abstract

**Background & Aim:** Liver fibrosis screening in primary care population is a major public health issue. The FIB-4 index is a simple non-invasive fibrosis test combining age, transaminases, platelets count, developed for the diagnosis of advanced fibrosis. The aim of our study was to evaluate the interest of liver fibrosis screening using systematic calculation of FIB-4 in routine blood analysis.

**Methods:** Between December 2018 and May 2019, we conducted a prospective screening of liver fibrosis in 134 158 patients during a medical check-up including routine blood analysis. Among these patients, 29 707 had transaminases and platelets counts available and benefited from an automatic calculation of FIB-4. Results were obtained from 21 French clinical laboratories in the Bouches du Rhône region.

**Results:** Among the 29 707 patients, 2160 (7.3%) had significant fibrosis (FIB-4>2.67). Individual investigation of patients with FIB-4>2.67 allowed to screen 1267 (1267/2160: 59%) patients who were not managed for any liver disease.

**Conclusions:** This work demonstrates the interest of FIB-4 for the screening of liver fibrosis in primary care population. Our study strongly supports this easy-to-implement strategy using a simple Fib-4 measure resulting from the use of available routine test results.

**Funding:** This research did not receive any specific grant from funding agencies in the public, commercial, or not-for-profit sectors.

**Clinical Registering Number:** MR-0314071019 (INDS: French National Institute for Medical Data)

## Introduction

Clinical laboratories report most test results as individual numerical or categorical values. However, individual test results are of limited diagnostic value [1]. To adequately use test results for patient diagnosis and management, clinicians must integrate many individual test results from a patient and interpret them in the context of clinical data and medical knowledge, judgment, and experience. However, this approach has some limitations. However, computational approaches can support the clinician, with the comprehension and interpretation of all the patient’s test results available. Therefore, it has the potential to enhance diagnostic values [2–5]. Many patients will have many of these individual test results, often spanning years.

Liver fibrosis screening in primary care population is a major public health issue. Among the causes of liver fibrosis, non-alcoholic fatty liver disease (NAFLD) is highly prevalent, affecting ∼25% of the population and is likely to increase further because of the obesity epidemic [6,7]. NAFLD is typically asymptomatic, and therefore most of the patients remain undiagnosed. The only available data in French primary care population reported that 2.6% of patients were identified with advanced liver fibrosis (Constances Cohort) [8]. Various scoring systems/tools are available for the non-invasive assessment of liver fibrosis. Many require measurement of the aspartate aminotransferase (AST), while others include transient elastography (Fibroscan) and serum fibrosis tests including serum enhanced liver fibrosis (ELF) test, Fibrotest, Fibrometer, Hepascore, or APRI [9–11]. Neither NICE nor EASL-EASD-EASO guidelines make specific recommendations regarding when to use liver biopsy in NAFLD assessment, although the EASL-EASD-EASO, APASL, AASLD guidance advocate the approach of applying non-invasive methods first, to avoid biopsies in low-risk cases [12–15]. In addition, Newsome et al. recommend the use of FIB-4 in patients with potential NAFLD for the management of abnormal liver enzymes [4].

The UK NAFLD survey attempted to capture data on which tools are currently most widely used for non-invasive fibrosis assessment [13]. The survey found that the AST is routinely measured in a hospital setting in 71.4% of cases, versus 33.9% of primary care cases. The survey indicates that primary care does not routinely perform any assessment of liver fibrosis, with only 7.9% routinely performing AST/ alanine aminotransferase (ALT) ratio in primary care. The most commonly used routine tests in secondary care patients with suspicion of liver disease are as follows: AST/ALT ratio (53%), transient elastography (Fibroscan) (50%), NAFLD fibrosis score (41%), FIB-4 score (16%), APRI score (6%), ELF test, or other serum fibrosis markers (5%).

Type 2 diabetes mellitus (T2D) is not only associated with NAFLD but also is an independent risk factor for the development of non-alcoholic steatohepatitis (NASH) [16–18]. Screening this high-risk population using a simple noninvasive tool is mandatory in order to prevent and avoid hepatic and extrahepatic worse manifestations[19].

The FIB-4 index is a simple non-invasive fibrosis test, combining age, transaminases, and platelet count, developed for the diagnosis of advanced fibrosis [20,21]. The score contributes to the assessment of NASH, hepatic C virus (HCV) or cholestatic, and metabolic liver diseases. There are four variables considered: patient’s age, AST, ALT, and platelet count. The FIB-4 equation is 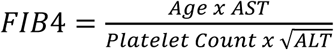. Cut-off values applied were >2.67 for severe fibrosis, and no fibrosis was defined as <1.30 in patients aged under 65 years and <2.00 for patients aged 65 years or higher [22,23].

The aim of our study was to evaluate the interest of liver fibrosis screening using systematic calculation of FIB-4 in routine blood analysis (during which a hepatic blood test check-up and platelet count were prescribed).

## Materials and Methods

Between December 2018 and May 2019, we conducted a prospective screening of liver fibrosis in 134 158 patients for whom routine blood tests were done during medical check-up addressed by primary care physicians. Fib-4 was systematically calculated in all patients for whom transaminases and platelets counts were available (ie. 29 707 patients).

Patients’ screening was performed using Microsoft Excel for Office 365 with Power Pivot solution (Microsoft, Redmond, WA).

Results were obtained from 21 French clinical laboratories located in Marseille, France. According to Article L1121-1 of the French Public Health law, non-interventional studies are not subjected to a legal framework. Non-interventional studies are defined as actions routinely performed without any additional procedure, unusual diagnostics, or monitoring. Patients were informed that biological results and samples could be used for research purposes and were free to refuse to participate. Study was submitted to French ethics committee (CPP Sud-Méditerranée II) and was registered as a reference methodology (MR-004) according to French regulations (registration number: 103140710192019, website: https://www.health-data-hub.fr/projets/etude-fib4-depistage-de-la-fibrose-hepatique-laide-du-fib-4-calcule-automatiquement-lors). Patients undergoing chemotherapy or with hematology disorders were excluded.

Age of patients, gender, platelet count, aspartate aminotransferase, alanine aminotransferase, and glycemia were retrieved from laboratory database. In case of multiple medical check-up of one individual patient, only the last one was considered for the analysis. All blood draws were performed under fasting conditions.

Two-sided Chi-square test was assessed to compare proportions at a significance level of 5%.

## Results

### Patients’ Characteristics

Among the 29 707 patients and according to FIB-4 fibrosis staging, 2160 (7.3%) had a higher risk of fibrosis (FIB-4>2.67), 22 529 (75.8%) had low risk of fibrosis (FIB-4<1.3 in patients under 65 years old and FIB-4<2 in patients aged over 65 years old), and 5018 (16.9%) had medium risk of fibrosis (**Figure 1**). A subgroup of 1267 patients (1267/2160: 59%) was identified and potentially not diagnosed or followed up for a liver disease (**Table 1**): patients were arbitrarily considered as managed for liver disease if they were addressed to the laboratory by a hepatogastroenterologist. Among the 2118 hyperglycemic patients (glycemia>7 mmol·L), 372 (17.6%) had FIB-4>2.67, 1323 (62.4%) had no-to-mild fibrosis, and 423 (20.0%) had undetermined fibrosis (Figure 2). The rate of significant fibrosis was higher in hyperglycemic patients compared with normoglycemic patients, 17,6 % [CI95 16.0-19.3%] vs. 8.6% [7.9-9.2%] (p<.0001), respectively.

**Table 1:** Characteristics of the patients

| Characteristics | All Patients<br>(n=29 707) | Fib 4 |  | Patients not managed<br>for liver disease<br>(n=1267) |
| --- | --- | --- | --- | --- |
|  |  | ≤2.67<br>(n=27 547) | >2.67<br>(n=2160) |  |
| Gender – N(%) |  |  |  |  |
| Male | 12525 (42%) | 11357 (41%) | 1168 (54%) | 684 (54%) |
| Female | 17182 (58%) | 16190 (59%) | 992 (46%) | 583 (46%) |
| Age – Mean (Sd) (years) | 54 (21) | 52 (20) | 76 (14) | 77 (13) |
| Platelet Count – Mean (Sd) (G/l) | 255 (81) | 262 (78) | 167 (65) | 161 (57) |
| Aspartate Aminotransferase – Mean (Sd) (IU/l) | 29 (63) | 25 (16) | 86 (218) | 60 (149) |
| Alanine Aminotransferase – Mean (Sd) (IU/l) | 28 (56) | 25 (31) | 61 (172) | 61 (172) |

**Figure 1:**
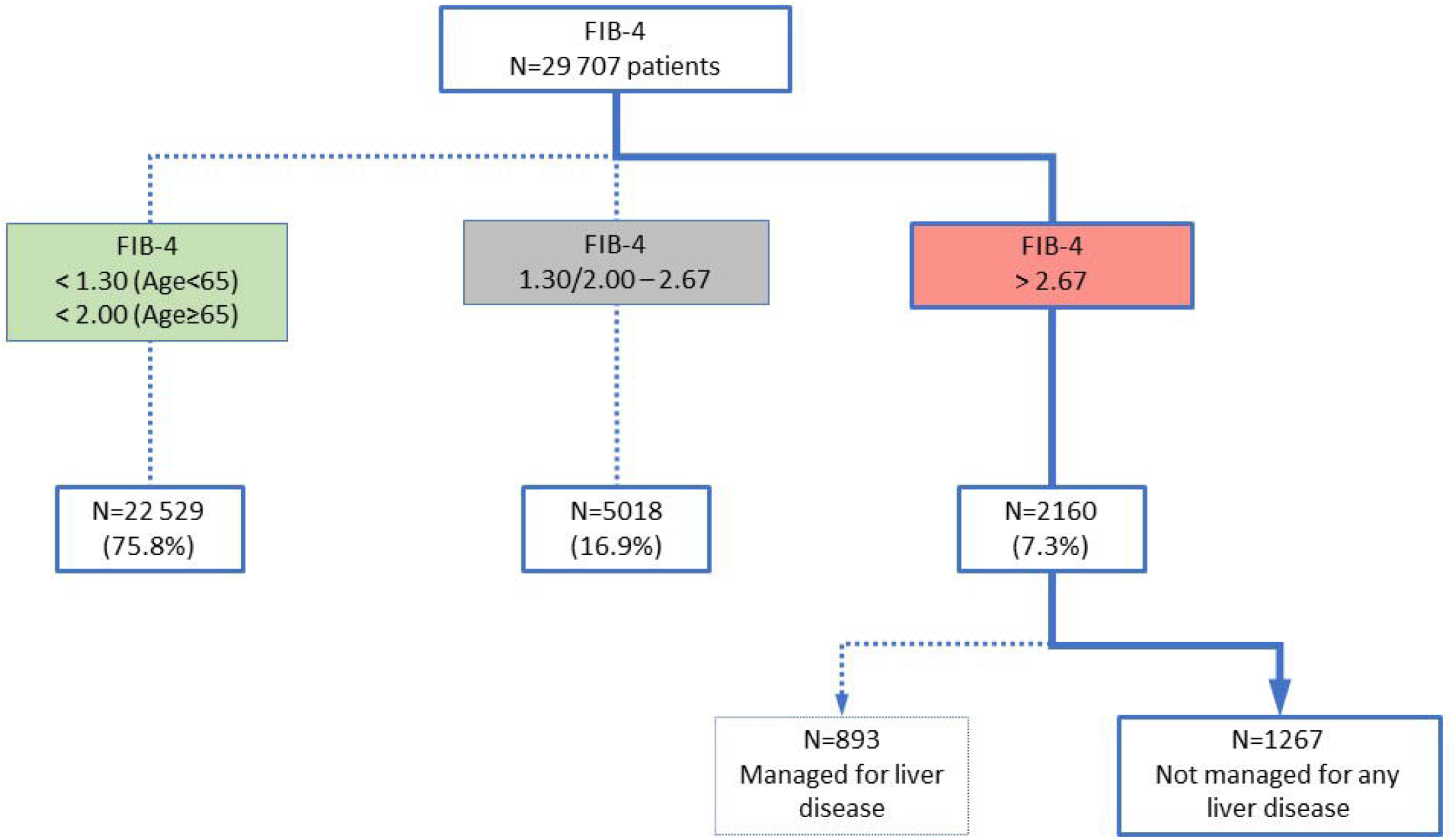
Study flowchart

**Figure 2:**
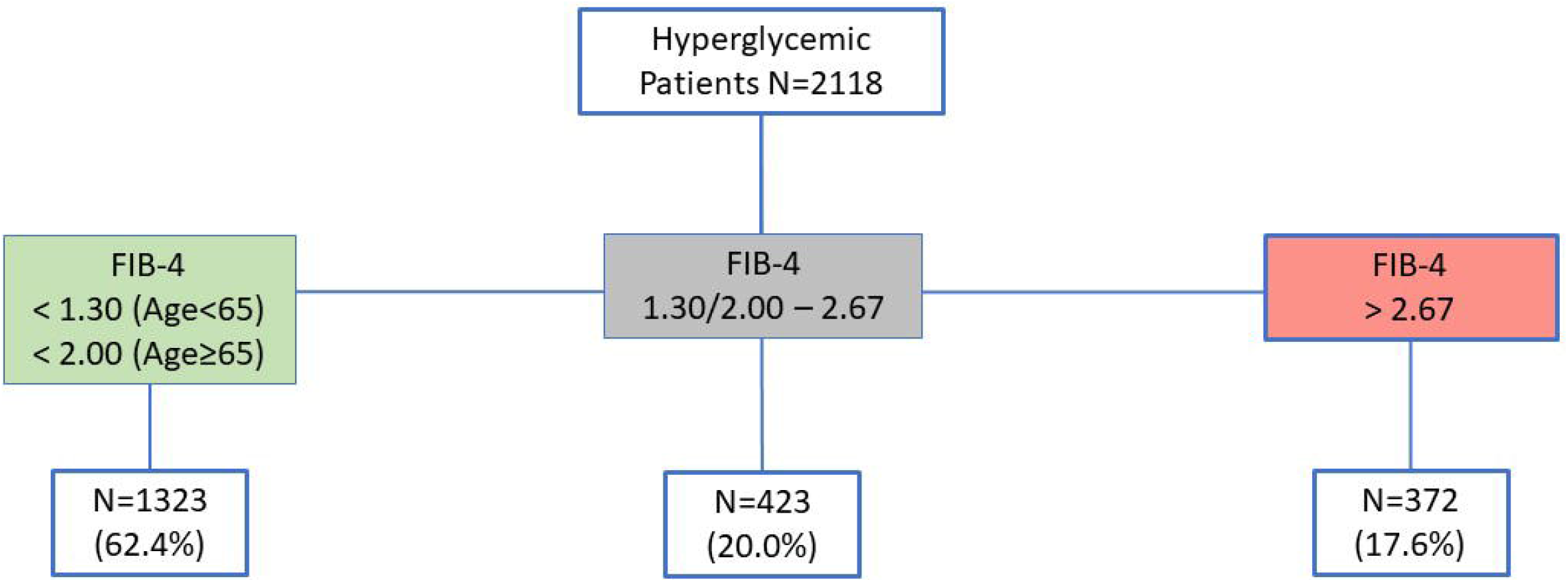
Flowchart of hyperglycemic patients

## Discussion

These findings highlight the substantial informational redundancy present in patients’ test results and offer a potential foundation for an innovative approach of clinical decision support aimed at integrating, interpreting, and enhancing the diagnostic value of a multi-analysis sets of clinical laboratory test results. This approach was used in the present study and allowed the identification of 1267 patients through a FIB-4 scoring above 2.67 and potentially not diagnosed and managed for liver disease (ie 59% of all patients with FIB-4 above 2.67). Our study provides an estimate of fibrosis prevalence in primary care practice and more importantly demonstrate the gap between patients with fibrosis who are undiagnosed and not managed for their liver disease and those who are diagnosed. This underscores the need for medical education on liver disease diagnosis and the utility of such tools to reinforce liver disease screening [5,24–28]. Two studies on primary care were performed in France: the first one (Constances) including 102 344 participants who were screened using the Fatty Liver index identified 16.7% of NAFLD patients; among them, 2.6 % using the FORNS index were identified with liver fibrosis [8]; the second one reported by Poynard et al. in individuals >40 years old in two French Social Security centers reported a presumed prevalence of advanced fibrosis (2.8%) and cirrhosis (0.3%)(not confirmed by liver biopsy)[29].

In hyperglycemic patients, the present study identified 17.6% with a Fib-4 > 2.67 among 2118 patients with a Fib-4 > 2.67, which was very similar to the study of Kwok R et al. (1918 patients with type 2 diabetes from Hong Kong), and the prevalence of increased liver stiffness (>9.6 kPa, suggestive of stage ≥F3) was 18% [30]. Liver enzymes are normal in up to 80% of NAFLD patients, and therefore cannot identify patients with a liver fibrosis [31].

A limit of this study is that no confirmation was obtained from patients regarding the management of their potential liver disease.

In conclusion, because the human brain faces great challenges in simultaneously considering a large number of data points, even the most experienced clinicians may be unable to extract all the useful information from existing clinical and laboratory data [3]. Electronic clinical decision support represents an important tool to improve test result interpretation and its efficiency for converting diagnostic data into useful information. Our study strongly supports this easy-to-implement strategy using a simple Fib-4 measure resulting from the use of available routine test results. That may represent an initial step in order to increase medical education and enhance fibrosis diagnosis.

## Data Availability

Data will be available immediately after publication with no end date to researchers who provide a methodologically sound proposal. Requests should be addressed by email to.

## Abbreviations

NAFLD: non-alcoholic fatty liver disease
AST: aspartate aminotransferase
ELF: enhanced liver fibrosis
ALT: alanine aminotransferase
Fibroscan: transient elastography
T2D: Type 2 diabetes mellitus
NASH: non-alcoholic steatohepatitis
HCV: hepatic C virus

